# Efficacy and safety of turmeric extracts in knee osteoarthritis: protocol for a systematic review and meta-analysis of randomised controlled trials

**DOI:** 10.1101/2020.05.06.20092726

**Authors:** Zhiqiang Wang, Ambrish Singh, Benny Antony

**Author notes:** Corresponding author: Dr Benny Antony*., Menzies Institute for Medical Research, University of Tasmania, 17 Liverpool St, Hobart TAS 7000, Tasmania, Australia. equal first authors (Zhiqiang Wang, Ambrish Singh).

## Abstract

Turmeric extracts have been used as a remedy for treating arthritis in traditional medicine. Recent years have witnessed the rise of different extracts from turmeric and randomised clinical trials (RCTs) evaluating the efficacy and safety of these extracts for the treatment of knee osteoarthritis (OA). This planned systematic review and meta-analysis aims to assess the efficacy and safety of turmeric extracts for the treatment of knee OA. Biomedical databases such as PubMed, Scopus, and Embase will be searched for RCTs reporting safety and efficacy of turmeric extracts for the treatment of knee OA. Cochrane risk of bias tool will be used to assess the methodological quality of the included studies, and a meta-analysis will be performed to pool the effect estimates.

## Background and rationale

Knee osteoarthritis (OA) is a common chronic disease that causes joint pain and knee function loss.^[1]^ There are currently no approved disease-modifying drugs available to treat OA. The pharmacological management options for OA are limited to analgesics, intra-articular corticosteroids, and NSAIDs.^[2]^ These medications only have a mild to moderate effect on pain, which results in patient dissatisfaction. Moreover, these drugs are associated with gastrointestinal, renal, and cardiovascular complications^[3]^ and are often contraindicated in patients with comorbidities.

Turmeric extracts, or *Curcuma longa* extracts, have been used as a remedy for treating arthritis in traditional medicine. Preclinical and clinical evidence suggests that *C. longa* extracts are effective and safe for the treatment of OA. The safety profile of turmeric makes it an ideal option for long-term treatment for OA patients who often have comorbidities. Recent years have witnessed the rise of different types of extracts from turmeric and randomised clinical trials (RCTs) evaluating the efficacy and safety of these extracts for the treatment of knee OA.

### Objectives

We aim to systematically review and meta-analyse the evidence from RCTs on efficacy and safety of turmeric extracts for knee OA. Efficacy will be evaluated in terms of improvement in symptoms (pain and physical function) and change in biomarkers (biochemical markers and imaging markers), and safety will be assessed from reported adverse events.

## Methodology

### Eligibility criteria

According to the criteria proposed by the PRISMA Extension Statement for Reporting of Systematic Reviews, the selection of studies will be based on the specific definition of PICOS items: Participants, Interventions, Comparators, Outcomes, and Study design.^[4]^

### Characteristics of participants (P)

The population of interest will include adult participants, at least 40 years old, of any sex and with a confirmed diagnosis of knee OA, possibly according to the criteria proposed by the American College of Rheumatology or similar.^[5]^ Moreover, studies involving mixed samples of patients with OA of the knee and/or hip may also be included, if found appropriate as per the inclusion criteria.

### Characteristics of interventions (I)

This study will focus on turmeric extracts from different *Curcuma* species (*C. longa, C. zedoaria, C. domestica*) and different enriched forms of extracts such as turmerosaccharides, curcuminoids, curcumin, jiang huang (Chinese name of *C. longa*), and such other formulations. RCTs evaluating combination therapy of turmeric as adjuvant or complementary to conventional drugs (e.g. paracetamol or NSAIDs) will be included, if the comparator group also received the same drugs. We will exclude trials with multi-herbal formulations. However, any turmeric extract formulation which includes benign bioavailability enhancing product will be included. For instance, piperine combined with turmeric will be included as it is a common additive to increase the bioavailability of curcumin.^[6]^

### Characteristics of comparators (C)

We will include the studies comparing turmeric extracts with active control (e.g. but not limited to, NSAIDs) or placebo for the treatment of knee OA.

### Characteristics of outcomes (O)

The studies reporting data for at least one of the following outcomes: pain, physical function, imaging biomarkers (x-ray joint space narrowing, MRI structural measures), biochemical markers, medication change, and adverse events will be included in the analysis.

For each outcome, data on baseline and follow-up values and/or mean change from baseline will be extracted. If the data are expressed in the graphical information, the numerical data will be extracted from graphs using the procedure (adapted) suggested by Guyot et al.^[7]^ If the studies do not provide complete data, authors of primary studies will be contacted through email to provide missing or additional data.

### Characteristics of study design (S)

Papers will be included if they used a randomised, quasi-randomised, controlled, blinded or non-blinded study design. Observational and non-randomised studies will be excluded.

## Primary and secondary outcomes

Our primary outcome will be change in knee pain. Secondary outcomes include change in physical function, change in biochemical markers, and change in imaging biomarkers.

## Information sources and search procedure

The search procedure will be implemented consistently with the following criteria.

### Electronic source and search strategy

Studies will be retrieved through a systematic search of the biomedical databases for the currently available literature on the use of turmeric extracts for knee OA. We will search online databases such as PubMed, Scopus, Embase, Web of Science, Cochrane Central Register of Controlled Trial, Google scholar, etc. from inception to May 2020. Both published and unpublished trials will be included, with the latter including, e.g. abstracts, conference proceedings and posters with available data. Studies that have compared interventions of interest and reported extractable data for at least one measure of pain, physical function, imaging biomarkers, biochemical markers, medication change, and adverse events will be included.

### Hand-searching

Abstract booklet from conference proceedings and poster sessions will be hand searched using the online sources of major international association involved in OA research: European League Against Rheumatism (EULAR), Osteoarthritis Research Society International (OARSI), American Academy of Orthopaedic Surgeons (AAOS), and American College of Rheumatology (ACR). Moreover, bibliography of the relevant meta-analysis and review articles will also be hand-searched and analysed for inclusion as per the eligibility criteria.

### Study selection

Study selection will be performed by two reviewers, independently, and each potential discrepancy will be discussed and resolved through consensus and/or consultation with senior authors.

### Assessment of risk of bias

The methodological quality of the included studies will be evaluated using the Cochrane risk of bias tool.^[8]^ Each article will be independently evaluated by two researchers using the Cochrane risk of bias tool. This double-check method will reduce the probability of an incorrect or inaccurate judgment of risk of bias. The Cochrane risk of bias tool considers characteristics of the following items: sequence generation, allocation concealment, blinding of participants, study personnel and outcome assessors, incomplete outcome data, selective outcome reporting, and other potential sources of bias. At the end of quality assessment, a consensus on final evaluation will be reached; any disagreements will be resolved by the discussion with senior authors.

### Data extraction

Two reviewers will extract data independently from the included studies for the following information: study design, characteristics of the population (age, sex, and BMI), sample size, intervention details and dosage, duration of follow-up, type of placebo or other control, outcome measurements, mean change values of the relevant outcome, the number of adverse events reported, and medication change. Intention-to-treat data will be used whenever available.

## Statistical analysis

The fixed-effect model will be used if included studies are homogeneous; otherwise, the random-effect model will be employed for the meta-analysis of both continuous and binary outcomes.^[9]^ The heterogeneity of the effect size across the trials will be tested using the Q statistics (P<0.05 was considered heterogeneous) and I^2^ statistic (I^2^ >50% will be considered heterogeneous).^[9]^ Due to different outcome measures, the change from baseline to follow-up scores will be translated into standard mean differences (SMD) using Hedges’ g effect sizes, as per the data availability. Statistical analysis will be performed using STATA version 16 (STATA Corp., Texas, USA) and Review Manager 5 (RevMan 5.3) (Copenhagen: The Nordic Cochrane Centre, The Cochrane Collaboration, 2014).

## Data Availability

Not applicable

